# Long-term Trend in Infant Mortality in India: A Joinpoint Regression Analysis for 1981-2018

**DOI:** 10.1101/2020.06.03.20120907

**Authors:** Aalok Ranjan Chaurasia

## Abstract

Infant mortality rate (IMR) in India remains high by international standards. India accounts for largest number of global infant deaths. This study analyses the trend in IMR in India over almost four decades beginning 1981 through 2018. The analysis is based on the official estimates of IMR available through sample registration system. Long-term trend in IMR is analysed by using joinpoint regression analysis which reveals that the trend in IMR in India changed four times during the period 1981-2018 and the trend has been different for different states of the country. The annual proportionate decrease in IMR was the most rapid during 1985-92 in the country and in most of its states but slowed down considerably during the period 1992-99. The significant deceleration in the decrease in IMR during 1992-99 appears to be the result of the policy shift in the delivery of maternal and child health services. Had the decrease in IMR not decelerated during 1992-99, the IMR in India would have been decreased to less than 20 by 2018.

## Introduction

The risk of death during the first years of life in India may be termed as very high by international standards. According to India’s official sample registration system, the infant mortality rate in the country during 2018 was 32 infants deaths for every 1000 live births (Government of India, 2020). On the other hand, according to the United Nations Inter-agency Group for Child Mortality Estimation (UN IGME) the infant mortality rate in India is estimated to be 29.9 infant deaths for every 1000 live births in the year 2018 (Unicef, 2019). Among the 192 countries of the world for which, estimates of infant mortality rate have been prepared by UN IGME, India ranks poor 139. UN IGME has also estimated almost 882 thousand infant deaths in India during 2018 which more than 18 per cent of the global infant deaths. This proportion is the highest among the 192 countries for which estimates are prepared by UN IGME. India’s National Health Policy 2017 has aimed at reducing the infant mortality rate in the country to 28 infant deaths for every 1000 live births by the year 2019 (Government of India, 2017).

Within India, infant mortality rate varies widely across states and Union Territories. According to India’s official sample registration system, the infant mortality rate in 2018 in the country varied from the lowest level of 4 infant deaths per 1000 live births in Nagaland to the highest level of 48 infant deaths for every 1000 live births (Government of India, 2020). Among the larger states of the country - states with a population of at least 20 million at the 2011 population census, the infant mortality rate in 2018 varies from 7 infant deaths per 1000 live births in Kerala to 48 infant deaths per 1000 live births in Madhya Pradesh. Among the 36 states and Union Territories of the country, the infant mortality rate in 2018 was less than 10 infant deaths per 1000 live births in six states and Union Territories but more than or equal to 40 infant deaths per 1000 live births in five states. There is no Union Territory where the infant mortality rate was 40 infant deaths per 1000 population or more in 2018 according to the sample registration system. The wide variation in the infant mortality rate across states/Union Territories of the country may be attributed to both wide variation in the level of infant mortality in the past and the variation in the historical trend the in infant mortality rate. For example, 20 years ago, in 1998, the infant mortality rate across 32 states/Union Territories of the country, as they existed at that time, varied from 16 infant deaths per 1000 live births in Kerala to 98 infant deaths per 1000 live births in Madhya Pradesh and Odisha according to the sample registration system (Government of India, 2000).

India’s official sample registration system provides annual estimates of infant mortality rate for the country as a whole and for its 15 states for the period 1981 through 2018. The time series of the infant mortality rate spanning almost four decades provides a unique opportunity to analyse the long term trend in the risk of death in the first year of life in the country and in its 15 states which account for almost 90 per cent population of the country according to the 2011 population census. Such an analysis, however, has never been carried out in India despite the availability of time series data on infant mortality rate for the country and for its 15 states from the official sample registration system. An analysis of the long term trend in the infant mortality rate is also useful in analysing the impact of different child survival interventions and programmes launched in the country from time to time. The history of child survival efforts in India may be traced back to 1978 when the Expanded Programme of Immunisation (EPI) was launched in the country. The EPI was followed by the Universal Immunisation Programme (UIP) in 1985 which was accorded the status of the National Technology Mission in 1986. In 1992, the UIP was replaced by the Child Survival and Safe Motherhood (CSSM) Programme and the Reproductive and Child Health (RCH) Programme in 1997. Subsequently, the RCH Programme became the lead programme of the National Rural Health Mission launched in 2005 (Chaurasia, 2017). The impact of these and many other child survival interventions on the pace of decrease in infant mortality rate in India has, however, never been analysed. It may be hypothesised that introduction of any new intervention or programme directed towards improving child survival should have resulted in accelerating the decrease in the infant mortality rate. If this hypothesis is true then the decrease in the infant mortality rate would have been accelerated after the introduction of the intervention or the programme.

In this paper, we identify the years when the trend in the infant mortality rate in the country and in its selected states have changed significantly during the period 1981 through 2018 or the joinpoints. Once the years when the trend in the infant mortality rate changed significantly or the joinpoints are identified, we calculate the annual percentage change in the infant mortality rate between two joinpoints to analyse how different child survival interventions and programmes have contributed to modifying the rate of decrease in the infant mortality rate. Such an analysis will help in assessing the effectiveness of different child survival interventions or programmes in accelerating the pace of decrease in the infant mortality rate in the country and in its constituent states.

The paper is organised as follows. The next section of the paper describes the data used in the analysis. The paper is based on the annual estimates of infant mortality rate available through India’s official sample registration system which is the only source of annual estimates of infant mortality rate in India. Section three of the paper outlines the methodology used for identifying the year(s) or time point(s) when the trend in the infant mortality rate has changed significantly. Results of the analysis are presented in section four. Section five discusses different child survival interventions and programmes that were introduced in the country from time to time to accelerate the decrease in the infant mortality rate and analyses how these interventions have contributed to the change in the trend in the infant mortality rate in the country and in its constituent states. The last section of the paper summarises the findings of the analysis and discusses its policy implications.

## Data Source

The analysis is based on the estimates of the infant mortality rate available through India’s official sample registration system which is a dual record system based on Chandra Sekar-Deming technique (Chandra Sekar and Deming, 1949) and now covers the entire country. The sample registration system is the only data source in the country which provides annual estimates of infant mortality rate for the country as a whole and for its constituent states and Union Territories. Below state - district - level estimates of infant mortality rate are, however, not available through the system. Estimates of the infant mortality rate are available through the system uninterruptedly for 48 years since 1971 which permits analysis of long-term trend in the infant mortality rate in the country. In the present analysis, estimates of infant mortality rate for the period 1981 through 2018 available through the system have been used. These estimates are available for the country as a whole as well as for 15 major states of the country - states with a population of at least 2 million at the 2011 population census - and all of these 15 states have been included in the present analysis.

Estimates based on the sample registration system are generally believed to be quite accurate, although, some under reporting of vital events has been reported in the system which varies from state to state. An investigation carried out way back in 1980-81 had revealed that around 3.1 per cent for the births were omitted by the system at the national level (Government of India, 1983). Another similar enquiry conducted in 1985 suggested that the omission rate had decreased to 1.8 per cent for births, although omission rates continued to vary from state to state (Government of India, 1988). On the other hand Mari Bhat (2002) has estimated that the sample registration system has missed about 7 per cent of the births but there has been no substantial change in the completeness. Recently, Yadav and Ram (2015) have estimated that, at the national level, 2 per cent births went unrecorded by the system during 1991-2000 and this proportion was 3 per cent during the period 2001-2011.

## Methods

We apply joinpoint regression analysis (Kim et al, 2000) to identify the year(s) or the time point(s) when the trend in infant mortality rate in India and in its 15 states for the period 1981 through 2018. Joinpoint regression analysis is used to study the trend that varies over time. It identifies the time point(s) at which the trend in the variable of interest significantly changes or the joinpoint(s), and then estimates the trend between two jointpoint(s) in terms of annual percentage change. The goal of joinpoint regression is not to provide the model that best fits the time series data. Rather, joinpoint regression provides the model that best summarises the trend in the data (Marrot, 2010).

The joinpoint regression model is different from the conventional piecewise or segmented regression model in the sense that the number of joinpoint(s) and their location(s) is estimated within the model and are not set arbitrarily as is the case with the piecewise or segmented regression. The minimum and the maximum number of joinpoint(s) are, however, set in advance but the final number of joinpoint(s) is determined statistically. The model identifies the year when there is a change in the trend and calculates the annual percentage change (APC) which is the change in rates between joinpoint(s). Using the APC, the annual average percentage change (AAPC) for the entire duration is calculated as the weighted average of APC of different segments with weights equal to the length of the segment. When there is no joinpoint or the number of joinpoint(s) is zero, the model reduces to simple linear regression model that fits a straight line to the time series data.

Let *y_i_* denotes the infant mortality rate for the year *t_i_* such that *t*_1_ < *t*_2_ <… <*t_n_*. Then the joinpoint regression model is defined as

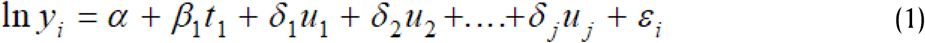

where

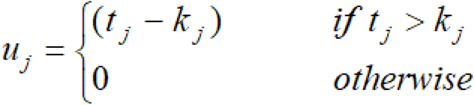

and *k_1_<k_2_*……<*k_j_* are joinpoints. The details of joinpoint regression analysis are given elsewhere (Kim et al, 2000; Kim et al, 2004).

Actual calculations were carried out using the Joinpoint Trend Analysis software developed by the Statistical Research and Application Branch of the National Cancer Institute of the United States of America. The software requires specification of minimum (0) and maximum number of joinpoints (>0) in advance. The programme starts with minimum number of joinpoints (0, which is a straight line) and tests whether more joinpoints are statistically significant and must be added to the model (up to the pre-specified maximum number of join points). The tests of significance is based on a Monte Carlo Permutation method (Kim et al, 2000).

We have used the grid search method (Lerman, 1980) which allows a joinpoint to occur exactly at time *t*. A grid is created for all possible positions of the joinpoint(s) or of the combination of joinpoint(s) and then the model is fitted for each possible position of the joinpoint(s) and that position of joinpoint(s) is selected which minimises the sum of squared errors (SSE) of the model. It may, however, be pointed out that even if the final selected model has *k* joinpoint(s), the slopes of the *k+1* temporal segments identified through the may not be statistically significant. The selection of *k* joinpoints simply implies that the model with these joinpoint(s) has a relatively better fit compared to all the other models within the pre-specified minimum and maximum number of joinpoints. When there is no joinpoint or the number of joinpoint(s) is zero, the model reduces to simple linear regression model that fits a straight line to the time series data.

Joinpoint regression analysis has frequently been used in the analysis of the trend in mortality and morbidity from specific causes (Tyczynski and Berkel, 2005; Doucet, Rochette and Hamel, 2016; John and Hanke, 2015; Akinyede and Soyemi, 2016; Mogos et al, 2016; Chatenoud et al, 2015; Missikpode et al, 2015; Rea et al, 2017; Qiu et al, 2008; Puzo, Qin and Mehlum, 2016). It has also been used for the estimation of population parameters under changing population structure (Gillis and Edwards, 2019). Chaurasia (2020) has used the segmented regression analysis to analyse the trend in economic growth in India.

In the present analysis, the year(s) or joinpoint(s) when the trend in the infant mortality rate has changed is matched with the time of introduction of any intervention or programme that is directed towards reducing the infant mortality rate. If the year(s) when the trend in the infant mortality rate has changed as obtained through the application of the joinpoint regression analysis matches with the year when a particular child survival intervention or programme was introduced, then, it can be argued that the child survival intervention or programme in question has been able to modify the trend in the infant mortality rate. A negative annual percentage change (APC) in the infant mortality rate obtained through the application of joinpoint regression analysis is an indication of the decrease in the infant mortality rate. A comparison of the APC before and after the inroduction of the child survival intervention or programme, therefore, provides the empirical evidence that the child survival programme or intervention in question has contributed to accelerating the decrease in the infant mortality rate. If there is no change in the trend in the infant mortality rate after the introduction of a particular child survival intervention or programme, then, it can be argued that there is no empirical evidence to suggest that the given child survival intervention or programme has contributed towards accelerating the decrease in the infant mortality rate.

### Infant Mortality in India 1981-2018

According to the sample registration system, the infant mortality rate in India was 110 infant deaths for every 1000 live births in 1981 which decreased to 32 infant deaths for every 1000 live births in 2018. Among the 15 states of the country, the infant mortality rate in 1981 was the highest in Uttar Pradesh followed by Madhya Pradesh and Odisha. In 2018, the infant mortality rate was the highest in Madhya Pradesh followed by Uttar Pradesh and Assam and Odisha. On the other hand, the infant mortality rate in 1981 was the lowest in Kerala followed by Karnataka and Maharashtra. In 2018, the infant mortality rate was again the lowest in Kerala followed by Tamil Nadu and Maharashtra. Kerala is the only state in the country, the rank of which in the infant mortality rate vis-a-vis other states of the country had not changed throughout the period 1981 through 2018. The inter-state coefficient of variation in the infant mortality rate increased from 0.284 in 1981 to 0.383 in 2018 indicating that there has been σ-divergence across the 15 states of the country which implies that the inter-state dispersion in the infant mortality rate has increased over time. This indicates an increase in the inequality in the risk of death during infancy across the states of the country.

The σ-divergence across states in the infant mortality rate has also been associated with the β-divergence. In a context of decreasing infant mortality rate, β-convergence in the infant mortality rate across states occurs when states with relatively high infant mortality rate have experienced a faster decrease in the infant mortality rate than regions with low infant mortality rate (Barro and Sala-i-Martin, 1992). This implies that the regression coefficient of the decrease in the infant mortality rate between time t=0 and t=1 on the infant mortality rate at time t=0 is negative. However, the regression coefficient of the decrease in the infant mortality rate between 1981 and 2018 in the 15 states of the country on the level of infant mortality rate in 1981 has been found to be positive, not negative. This means that there has been β-divergence across the states of the country during the period under reference as far as the decrease in the infant mortality rate is concerned. This essentially implies that the decrease in the infant mortality rate during 1981-2018 has been relatively faster in those states where the infant mortality rate was relatively low in 1981 as compared to the states where the infant mortality rate was relatively high in 1981. It has been argued that when the infant mortality rate decreases, more and more infant deaths gets concentrated in what is known as the hard rock of infant mortality so that it is relatively easier to reduce infant mortality when the infant mortality rate is high than when the infant mortality rate is low. However, such a trend is not visible across the states of the country rate. The decrease in the infant mortality rate appears to be relatively slow in those states of the country where the infant mortality rate was relatively high in 1981.

### Joinpoint Regression Analysis

Results of the application of joinpoint regression to the time series data of infant mortality rate in India and states for the period 1981 through 2018 are presented in figure 1. Two observations are very much apparent from the figure. First, there has been frequent change in the trend in the infant mortality rate in the country and in most of its states during the period under reference. At the national level, the trend in the infant mortality rate changed significantly four times during 1981 through 2018 so that the annual percentage change (APC) was different in different time periods. In other words, the decrease in infant mortality rate in the country during the period 1981 through 2018 has not been uniform. The annual percentage decrease in infant mortality rate in the country more than doubled during the period 1988 through 1991 as compared to the period 1981 through 1988 but the decreased slowed down significantly during the period 1991 through 1999 and accelerated during the period 1999 through 2007. Although the decrease accelerated further during the period 2007 through 2018, yet, the annual percentage decrease in the infant mortality rate was the highest during the short duration 1988 through 1991.

**Figure 1.**
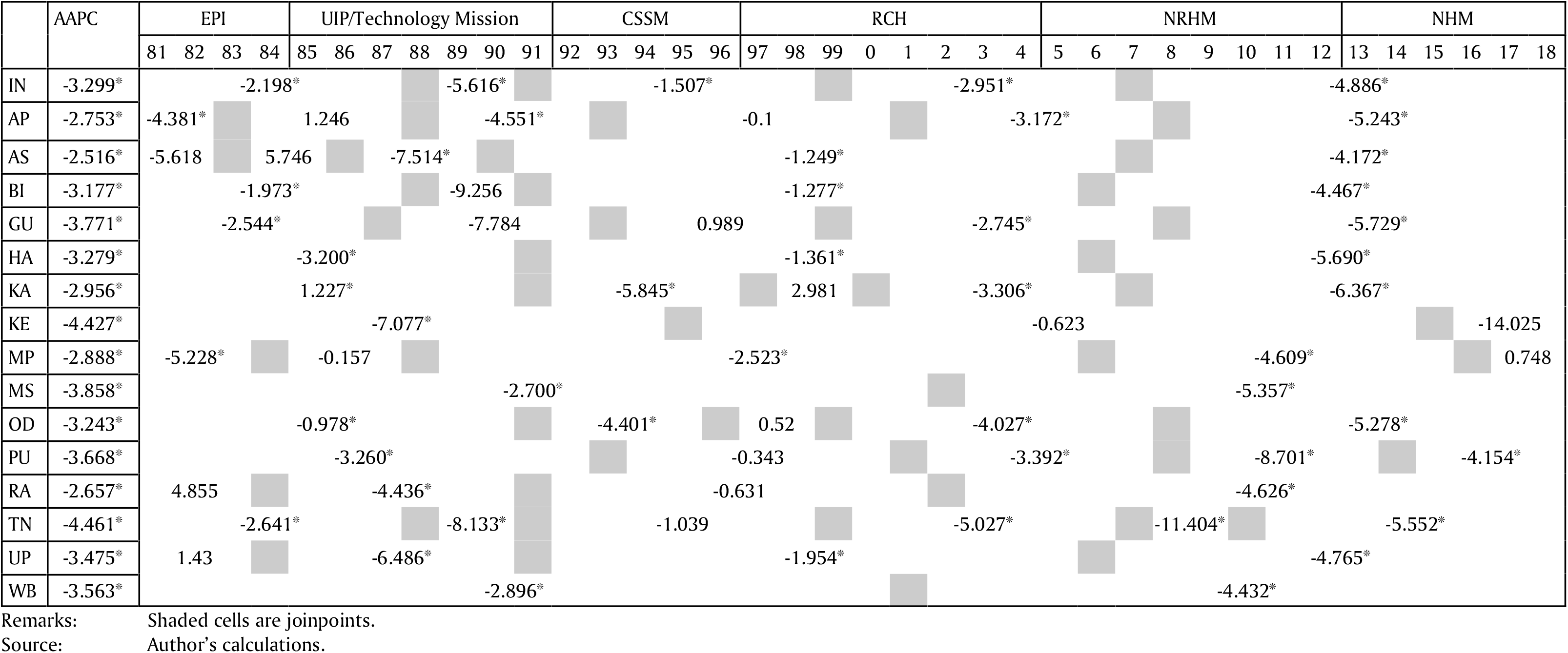
Results of the joinpoint regression analysis (APC in different periods of the duration 1981-2018), India and states.

It would have been interesting to project the infant mortality rate in the year 2018 under differing trends in the infant mortality rate revealed through the present analysis. For example, if the APC during 1981-88 would have been maintained during the period 1988 through 2018, the infant mortality rate in the country would have decreased to around 48 infant deaths per 1000 live births by the year 2018 (Figure 2). On the other hand, if the APC during 1988-92 would have been maintained during the post 1992 period, the infant mortality rate would have decreased to only 16 infant deaths per 1000 live births by the year 2018. By contrast, if the APC during 1992-99 would have been maintained during the post 1999 period, the infant mortality rate in the country would have decreased to only about 52 infant deaths per 1000 live births in 2018. The decrease in infant mortality rate accelerated during 1999-2007. If the APC during 1999-2007 would have been maintained after 2007, the infant mortality rate in the country would have decreased to around 39 infant deaths per 1000 live births in 2018. However, the decrease in the infant mortality rate accelerated further during the period 2007-2018 so that the infant mortality rate actually decreased to 32 infant deaths for every 1000 live births in 2018. It is obvious from the figure 2 that a significant slow down in the decrease in the infant mortality rate in the country during the period 1992 through 1999 as revealed through the present analysis has primarily been responsible for the observed in the infant mortality rate in the country. The slow down in the decrease in the infant mortality rate during 1991-99 appears to have delayed considerably the progress towards the reduction in the infant mortality rate in the country.

**Figure 2.**
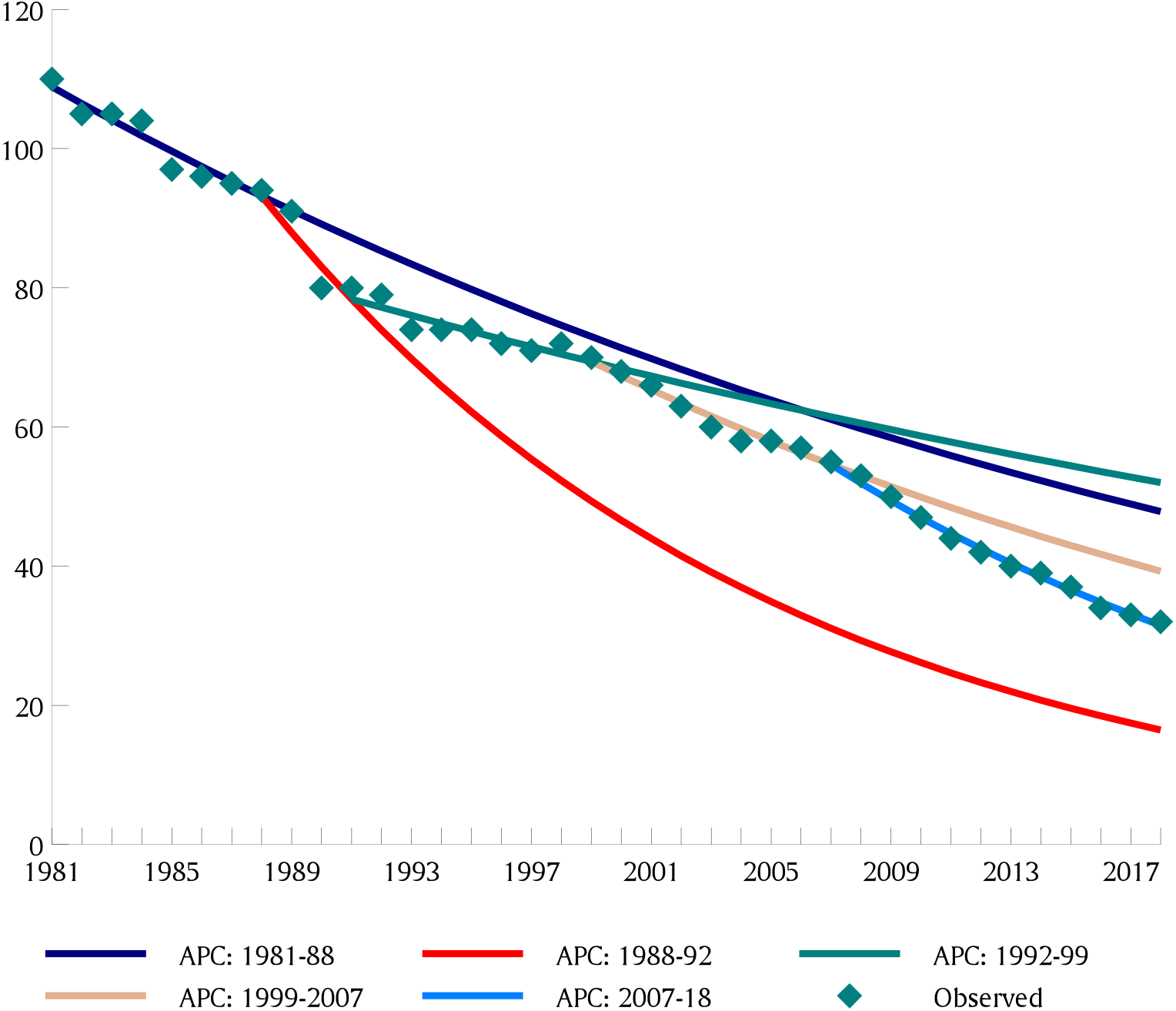
Observed trend in infant mortality rate in India and projected trend according to the annual percentage change in different time periods.

Figure 1 also suggests that there is no state in the country where the decrease in the infant mortality rate has been linear during the period under reference or the number of joinpoints is zero. However, the number of joinpoints varies from only one in Maharashtra and West Bengal to five in Andhra Pradesh and Tamil Nadu, reflecting the complex nature of the trend in the infant mortality rate in the states of the country. Figure 1 also suggests that the joinpoints or the years when the trend in the infant mortality rate changed significantly have also been in general different in different states. In other words, the trend in the infant mortality rate during the 38 years between 1981 and 2018 has essentially been different in different states of the country. This implies that of the country the trend in the infant mortality rate in different states of the country has been influenced by state-specific factors also.

The joinpoint regression analysis also suggests that the pace of decrease in the infant mortality rate or the annual percentage change (APC) has been different in different time periods in all states. For example, in five states - Assam, Bihar, Gujarat, Rajasthan and Uttar Pradesh - the annual percentage decrease in the infant mortality rate was the most rapid around the period 1985-91 when the Universal Immunisation Programme/National Technology Mission on Immunisation was launched. Had the annual percentage decrease in the infant mortality rate achieved during this period in these states would have been maintained after 1991, then there would have been a very significant reduction in the infant mortality rate in these states by the year 2018. However, in all these states, the decrease in the infant mortality rate either slowed down very significantly or the infant mortality rate increased instead decreased during the period when the Child Survival and Safe Motherhood (CSSM) Programme and Reproductive and Child Health (RCH) Programme was implemented in the country. It was only after the launch of the National Rural Health Mission that the decrease in the infant mortality rate in these states accelerated but the decrease has not been large enough to compensate for the very substantial slowdown in the decrease in infant mortality rate during the period of implementation of CSSM and RCH Programmes.

The case of Madhya Pradesh is typical. This state recorded the fastest decrease in the infant mortality rate during the period 1981-84 but the decrease in the infant mortality rate virtually stagnated during the early years of the Universal Immunisation Programme/National Technology Mission and picked up to some extent only after 1988. It was only after the launch of the National Rural Health Mission that there has been a modest acceleration in the decrease in the infant mortality rate in the state but the trend appears to have reversed after 2016 so that the infant mortality rate increased instead decreased during the period 2016-18. Madhya Pradesh is the only state in the country where the trend in the infant mortality rate reversed in recent years. The state has the dubious distinction of having the highest infant mortality rate amongst the states and Union Territories of the country since 2004 according to the official sample registration system. There is however little evidence to suggest that the pace of the decrease in the infant mortality rate in the state is accelerating.

The pace of the decrease in the infant mortality rate during 1981 through 2018 in different states of the country may be captured through the average annual percentage change (AAPC) in the infant mortality rate as obtained from the joinpoint regression analysis. The AAPC during 1981-2018 has been the highest in Tamil Nadu closely followed by Kerala, the only two states in the country where AAPC was more than 4 per cent during the period under reference. On the other hand, the AAPC was the lowest in Assam. Other states where AAPC during 1981-2018 was less than 3 per cent are Andhra Pradesh, Karnataka, Madhya Pradesh and Rajasthan. Among the four states, Madhya Pradesh and Rajasthan are the high infant mortality states whereas the infant mortality rate in Andhra Pradesh and Karnataka is relatively low.

The variation in the pace of decrease in the infant mortality rate across 15 states as reflected through AAPC suggests that the effectiveness of child survival interventions or programmes in reducing the infant mortality rate has varied widely across states. However, very little is currently known about the reasons behind the inter-state variation in the effectiveness of child survival interventions and programmes in accelerating the pace of decrease in the infant mortality rate. AAPC is determined by APC in different time intervals of the period 1981-2018 with the length of the time interval as weight. This means that the APC in a given time interval reflects the effectiveness of the child survival intervention being implemented in that time interval. According to this perspective, the effectiveness of the Child Survival and Safe Motherhood (CSSM) Programme and the Reproductive and Child Health (RCH) Programme in accelerating the decrease in the infant mortality rate appears to be poor in the country and in its most of the states because the annual percentage decrease in the infant mortality rate slowed down considerably during the period when these programmes were being implemented in the country. Ideally, these programmes should have an incremental effect on the APC but, as may be seen from figure 1, this has not happened in the country and in most of its states so that the decrease in the infant mortality rate slowed down considerably during the period when these programmes were implemented. Had these programmes were able to accelerate the decrease in the infant mortality rate, the pace of decrease in the infant mortality rate during the period 1981 through 2018 would have been substantially more rapid. The decelerated decrease or even the increase in the infant mortality rate during the period of implementation of these programmes appears to be primarily responsible for the deceleration of the decrease in the infant mortality rate in the country and in most of its states.

## Discussions and Conclusions

The infant mortality rate in India remains high by international standards and the country accounts for the largest proportion of the global infant deaths according to the estimates prepared by UN IGME (Unicef, 2019). This is so when the infant mortality rate in the country has decreased over the last 50 years according to India’s official sample registration system. The present analysis, however, indicates that decrease in the infant mortality rate in the country and in many of its states has not been consistent during the period 1981 through 2018 and there had been a very significant deceleration in the decrease in the infant mortality rate during the period 1992 through 1999. If the decrease in the infant mortality rate would have not decelerated during 1992-99, the gain in terms of the improvement in the risk of death in the first year of life in India would have been significantly more respectable in the international context. The same would have happened in most of the high infant mortality states of the country. It appears that India and many states of the country missed the opportunity of rapid improvement in the probability of survival in the first year of life that was created during the period 1988-92. A significantly decelerated decrease in the infant mortality rate during the period 1992-99 appears to be responsible for this loss of opportunity. The decrease in the infant mortality rate accelerated after 1999 but the acceleration was not substantial enough to compensate for the loss accrued due to a decelerated decrease in the infant mortality rate during the period 1992-99.

The deceleration in the decrease in the infant mortality rate in the country after 1992 appears to be the result of a major shift in the official policy towards meeting the health needs of women and children. During the period 1985 through 1992, the focus of the delivery of maternal and child health services in the country was on child immunisation against vaccine preventable diseases through the Universal Immunisation Programme (UIP) which was given the status of National Technology Mission in 1986. However, under the Child Survival and Safe Motherhood Programme launched in 1992, the focus of maternal and child health activities in the country shifted to preventing maternal deaths through emergency obstetric care. The rationale behind this shift appeared to be a report prepared by the World Bank and UNICEF which estimated that about 0.5 million women died every year in India as the result of complications during pregnancy and child birth (Mathur and Reddy, 2019). Another concern was near stagnation in the decrease in the perinatal and neonatal mortality (Ghosh, 1987). Although, child survival remained an integral component of the Child Survival and Safe Motherhood Programme, yet the focus of the official efforts in the delivery of maternal and child health services was distinctively upon preventing maternal deaths through strengthening and expanding emergency obstetric care services. It was also conjectured that the strengthening and expanding emergency obstetric care services would also contribute to the prevention of neonatal and perinatal deaths leading to accelerated decrease in neonatal mortality rate which will contribute to the decrease in the infant mortality rate. The present analysis, however, suggests that the decrease in the infant mortality rate decelerated, instead accelerated, in the country and in majority of its states after this policy shift in the delivery of maternal and child health services. The focus on safe motherhood interventions appears to have actually resulted in a neglect of child survival interventions after 1992.

The residual attention accorded to child survival interventions under the CSSM Programme may be judged from the fact that the proportion of children aged 12-23 months who were fully immunised increased from around 35 per cent in 1992-93 to only around 42 per cent in 1998-99 - an increase of only around 6 percentage points over a period of six years - according to the National Family Health Survey (IIPS, 1995; IIPS and ORC Macro, 2000). Although, efforts were also made under the CSSM Programme to reinforce the Oral Rehydration Therapy (ORT) and Acute Respiratory Infection (ARI) Programmes for control of diarrhea and pneumonia in children, yet the preoccupation with strengthening and expanding emergency obstetric care services to prevent maternal deaths put the implementation of different child survival interventions in a residual environment.

The residual attention given to child survival interventions, especially, child immunisation appears to have continued even after the CSSM Programme was expanded into the RCH Programme in 1996. According to the National Family Health Survey 2005-06, the full immunisation coverage rate in the country increased by just 1.5 percentage points between 1995-96 and 2005-06 (IIPS and Macro International, 2007). It was only after the launch of the National Rural Health Mission in 2005 that there had been an accelerated increase in the proportion of fully immunised children in the country (Figure 3). However, even after almost five decades of the launch of the Expanded Programme of Immunisation in the country, the cherished goal of universal immunisation of all infant still remains illusive in the country. According to the National Family Health Survey 2015-16, only 62 per cent of the children aged 12-23 months were found to be fully immunised and this proportion was less than 54 per cent in case of full immunisation coverage by 12 months of age (IIPS and ICF, 2017). Had India been able to maintain the child survival momentum generated during the period of Universal Immunisation Programme, the infant mortality rate in India would have decreased to less than 20 infant deaths per 1000 live births against the prevailing infant mortality rate of 32 infant deaths per 1000 live births and many of the high infant mortality rate states of the country would have been able to drastically reduce the infant mortality rate, particularly, because of a decelerated decrease in the infant mortality rate during the period 1992-99. Although, reasons for the decelerated decrease in the infant mortality rate in the country and in many of its high infant mortality rate during the period 1992-99 have never been explored, yet, it appears that a shift in the basic approach of maternal and child health services from child survival to safe motherhood appears to be responsible for the deceleration in the decrease in the infant mortality rate.

**Figure 3.**
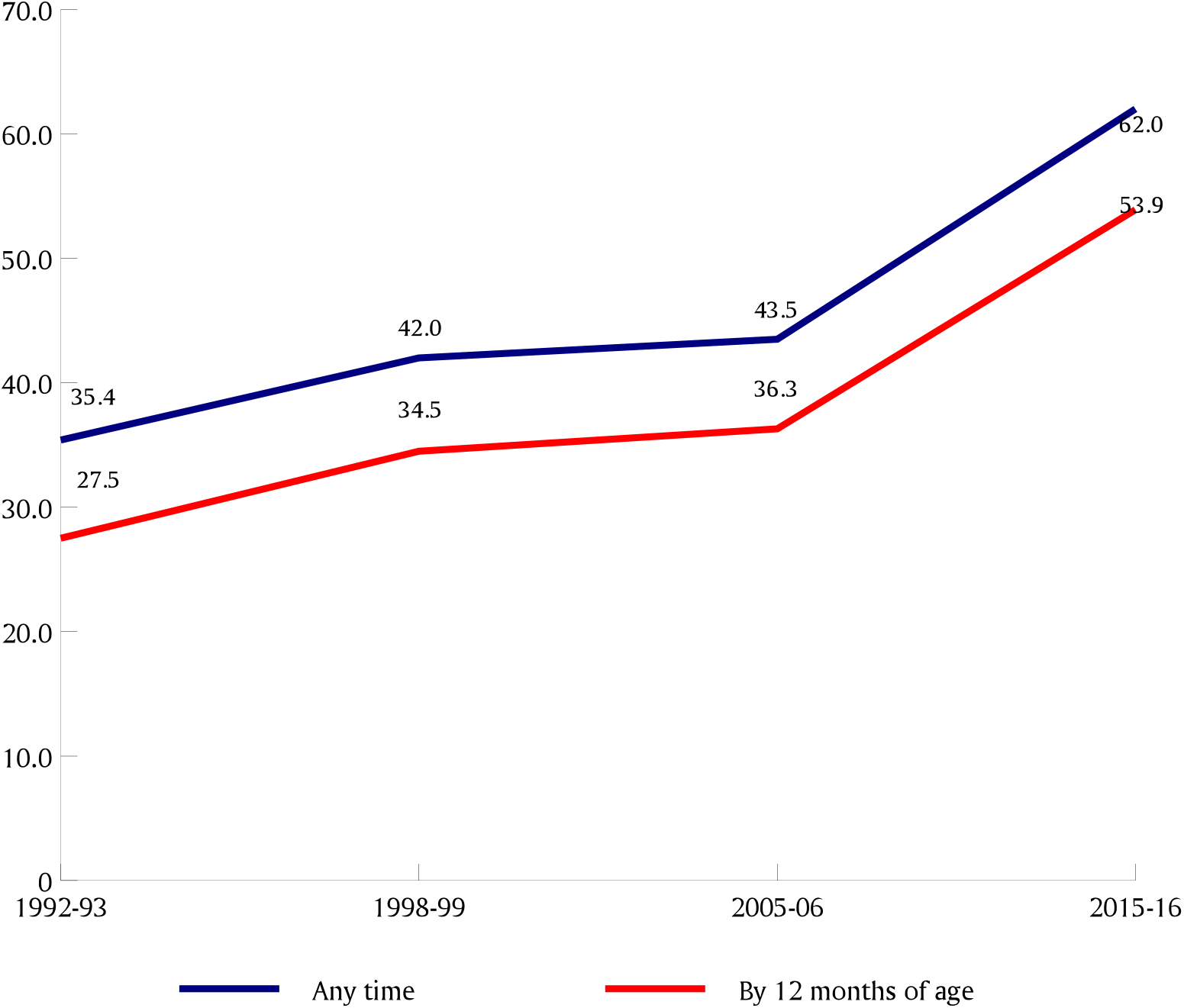
Trend in the proportion of children aged 12-23 months who are fully immunised.

The introduction of CSSM Programme in 1992 also marked a shift in the basic approach to maternal and child health services from the public health approach to risk-based institutional care to prevent premature deaths. The public health approach is essentially directed towards universalisation of the key child survival interventions such as immunisation against vaccine preventable diseases, oral rehydration therapy to prevent deaths due to dehydration during diarrhoea, home-based management of acute respiratory infections and a behavioural practices, especially those related to infant feeding. Because of the focus on universalisation, the foundation of public health is built upon the extension approach which emphasises reaching the community. The risk-based institutional care, on the other hand, calls for accessing a facility whenever there is a risk of a adverse health outcome. It is argued that in the midst of such shift in the approach to the delivery of maternal and child health care services, the health and well-being of children can easily slip from the view (National Research Council, 1996). This appears to have actually happened in India.

## Data Availability

Data used in the analysis are official data and is available at www.censusindia.gov.in

